# Alteration of serum leptin and LEP/LEPR promoter methylation in Prader-Willi syndrome

**DOI:** 10.1101/2021.12.15.21267839

**Authors:** Jelte Wieting, Kirsten Jahn, Vanessa Buchholz, Ralf Lichtinghagen, Stefan Bleich, Christian K. Eberlein, Maximilian Deest, Helge Frieling

## Abstract

Prader-Willi syndrome (PWS) is a rare neurodevelopmental disorder based on a loss of paternally expressed but maternally imprinted genes in chromosome region 15q11-13. During child development, PWS usually results in insatiable appetite with subsequent obesity representing the major mortality factor. The neurobiological basis of PWS-typical hyperphagia has remained poorly understood. Many PWS-typical abnormalities are based on hypothalamic dysregulation, the region in which hunger and satiety are hormonally regulated, with the hormone leptin being a main long-term regulator of satiety. Previous studies in PWS have inconsistently shown leptin alterations solely in early childhood, without investigating the leptin system on an epigenetic level.

The present study investigates serum leptin levels (S-leptin) and methylation of the leptin (LEP) and leptin receptor gene (LEPR) promoter in 24 individuals with PWS compared to 13 healthy controls matched for sex, age, and body mass index (BMI) and relates the results to the extent of hyperphagia in PWS.

S-Leptin levels were obtained by Enzyme-linked Immunosorbent Assay. LEP/LEPR-promoter methylation was assessed by DNA-bisulfite-sequencing, hyperphagia by Hyperphagia Questionnaire for Clinical Trials (HQ-CT). PWS and control groups differed significantly in S-leptin levels with higher S-leptin in PWS. Methylation analysis showed significant differences in mean promoter methylation rate both for LEP and LEPR with a lower methylation rate in PWS. LEPR, but not LEP methylation correlated with S-leptin levels. S-leptin and both LEP and LEPR methylation did not correlate with HQ-CT scores in PWS.

The present study is the first to show significantly elevated S-leptin levels in an adult PWS cohort combined with an altered, downregulated LEP and LEPR promoter methylation status compared to BMI-matched controls. Analogous to previous studies, no link to the behavioral dimension could be drawn. Overall, the results suggest an increased leptin dysregulation in PWS, whereby the findings partly mirror leptin resistance seen in non-syndromic obesity.

## INTRODUCTION

Prader-Willi syndrome (PWS) is a rare neuronal developmental disorders with an estimated prevalence of 1:15000 (1). The syndrome is genetically determined by a loss of normally paternally expressed but maternally imprinted genes on chromosome 15q11-13. Essentially, three different genetic subtypes can lead to PWS. Most common, in about 70% of cases, is a large deletion of the paternal allele of chr15q11-13 (delPWS). In about 25- 30% of cases a maternal uniparental disomy of chromosome 15 (UPD) and in rare cases, a defect of the imprinting center (IC) occurs (2).

Newborns initially present with a failure to thrive and muscular hypotension. During infancy, PWS individuals rapidly develop an insatiable appetite leading to massive weight gain unless food intake is regulated. Individuals with PWS show a deficiency of growth hormones and hypogonadism, accompanied by a dysregulation of body temperature and altered pain tolerance, which can essentially be explained by hypothalamic dysfunction (3). In addition, in almost all cases there is a mild to moderate intellectual impairment as well as an overall delay in language and motor development. But also, individuals with PWS exhibit a certain behavioral phenotype characterized by temper outbursts, repetitive and ritualistic behaviors, and skin-picking(4). Despite improved diagnostics and therapy, life expectancy is still assumed to be reduced in people with PWS. Obesity and its sequelae such as cardiovascular diseases, diabetes, as well as sleep apnea and other respiratory complications are the major driving factor for the increased mortality rate in people with PWS (5). Therefore, basic research into the causes of hyperphagia and obesity in PWS is of particular importance for improved diagnosis and treatment.

In human, satiety is induced essentially via stimulation of melanocortin-4 receptors (MCR-4) by the proopiomelanocortin (POMC) derivative alpha- melanocyte stimulating hormone (alpha-MSH) in the paraventricular nucleus of the hypothalamus. Besides stimulation of POMC, leptin inhibits AgRP- and neuropeptide Y (NPY)-producing neurons in the arcuate nucleus of hypothalamus, hormones, which have an appetite-increasing effect via MCR-4 inhibition. Leptin has an appetite suppressing, energy expenditure increasing and consequently body weight reducing effect (6). Animal studies suggest that leptin receptors are also found in the ventral tegmental and other midbrain areas, in which the regulation of the endogenous reward system is mediated (7).

In humans, the hormone leptin is encoded on the LEP gene located on chromosome 7q31.3 and composed of three exons and two introns. It is largely expressed in adipocytes, but also to a lesser extent centrally in the hypothalamus and pituitary gland, as well as other tissues (8). The physiological regulatory mechanisms behind leptin expression are complex. In healthy individuals, leptin blood levels correlate with body fat stores (9). In women, circulating leptin is significantly elevated compared to men. With increasing age leptin levels decrease (10). Fasting causes a decrease in leptin levels in humans, whereas levels re-increase after days of refeeding (11). It is important to note that leptin levels do not increase in response to individual meals (12). Leptin can be considered as a long-term indicator of the body’s energy reserves.

Leptin exerts its effect via leptin receptors. The LEPR gene is located on chromosome 1 (1p31) and consists of 18 exons and 17 introns. One of the splice variants of the LEPR gene, the longest isoform LEPRb is most widely expressed in the human brain, particularly in the hypothalamus and cerebellum, although leptin receptors are also expressed in other tissues such as the vasculature and stomach. Only the long isoform LepRb contains the intracellular motif required for leptin-mediated POMC-activation in the hypothalamus, making this isoform the main mediator of the above-mentioned leptin-specific effects (13).

Due to its size, leptin is unable to cross the blood-brain barrier (BBB) by diffusion. The neurobiological mechanisms of transporting leptin across the BBB are mainly unclear. In obese individuals, elevated leptin levels continue to be seen in both blood and adipocytes, but no significant leptin effect can be discerned. Accordingly, leptin resistance in obesity is assumed. Most of the evidence on the cellular and molecular mechanisms of leptin resistance has been obtained in experimental rodent models. Obese mice showed resistance to peripherally administered leptin but responded to intracerebroventricular infusions of leptin. The efficiency of central leptin uptake measured as CSF/plasma leptin ratio was 4.3-fold higher in lean than in obese subjects indicating a dysregulated leptin transport across the BBB to be associated with leptin resistance.

The causes for obesity in PWS are currently understood in rudimentary terms at best. Misbalances in the abovementioned hypothalamic satiety center and the various hormones influencing it have been suspected with the hyperphagia typical of PWS. Regarding leptin, significant differences in blood levels compared with obese controls have so far only been demonstrated for young children with PWS. Previous studies showed significantly higher leptin levels in infants <6 years of age compared to weight-matched controls in collectives sized 42 PWS to nine control (14) and 33 PWS to 28 control subjects (15), whereas this finding was absent in children >6 years of age in a study sized 35 PWS (n=21 <6y, n=14 >6y) and 31 age and BMI-matched controls (16). 22 adult patients with PWS showed elevated blood leptin levels compared to 54 healthy controls (17), but in adult PWS there were no significant differences compared to weight-adapted control subjects in cohorts of 20 PWS to 14 control (18) and 28 PWS to 29 obese control subjects (19). An older study by Butler et al. also showed no relevant differences between PWS and BMI-matched controls (PWS n=19 obese and n=14 non- obese, controls n=28 obese and n=16 non-obese), with the exception that non-overweight male PWS subjects had five times higher plasma leptin levels than controls (20). On a genetic level, Lindgren et al. examined leptin mRNA levels in subcutaneous adipose tissue in comparison of healthy controls and PWS children on the one hand and non-syndromic obesity on the other. Leptin mRNA levels were found to be significantly increased in both PWS and non-syndromic obese controls compared with healthy subjects, but there was no significant difference between PWS and non-syndromic obese controls. Both leptin serum and leptin mRNA levels correlated with BMI in PWS. However, the results are clearly limited by the small number of cases (n=6 each) (21).

Preliminary studies illuminated the influence of epigenetic mechanisms on hormone balance, including leptin, which is investigated in the present work. One important epigenetic mechanism is the methylation of cytosine bases in the DNA strand, which occurs only in the context of cytosine-guanine dinucleotides (CpGs). Methylation of the gene promoter is of particular interest in that it represents an essential regulatory unit for gene expression. Among other mechanisms, increased methylation impedes the binding of transcription factors so that protein biosynthesis is subsequently suppressed (22).

In the context of metabolic diseases, research results are available from both animal and human studies regarding altered LEP methylation status and leptin expression. In human studies, an association between decreased LEP-methylation status and obesity has been demonstrated in male children (23). In obese adolescents, a decreased methylation status of the LEP promoter region was detected in peripheral blood (24). Overall, the data predominantly indicate hypomethylation of the LEP promoter region in metabolic disorders. In general, an inverse correlation between LEP promoter methylation and leptin expression is assumed (25). In our previous publication on the subject, we examined 98 obese patients before and after bariatric surgery with respect to LEP promoter methylation and leptin expression. Consistent with previous studies, we saw increased leptin serum levels before bariatric surgery. But in contrast to the majority of studies regarding LEP promoter methylation, we found an increased LEP promoter methylation before surgery compared with postoperative levels (26).

Alterations of the LEPR gene in obesity have been less studied overall. In the aforementioned study on bariatric surgery, there was an increased postoperative LEPR methylation accompanied by an inverse association between LEPR methylation and S-leptin levels. Furthermore, another study on the influence of pre-pregnancy BMI on gestational weight suggested an inverse correlation between LEPR methylation and LepRb expression (27).

In the present study, we investigate the leptin system in an adult PWS collective compared to healthy BMI-matched controls. For the first time, we assess the regulatory level in terms of LEP and LEPR promoter methylation in PWS. The results will be related to the extent of hyperphagia the PWS subjects undergo, which is assessed by a specific questionnaire.

## METHODS

### STUDY CHARACTERISTICS

The study included 24 subjects with PWS (14 male, 10 female) and 13 healthy control subjects (8 male, 5 female) matched for sex, age, and body mass index (BMI). Control subjects with a difference of +/- 5 years and +/- 1 kg/m^2^ BMI compared to the study cohort were considered. The control subjects were recruited via public tenders analogously and online. The healthy control subjects confirmed during the interview that they had not been diagnosed with any mental illness at the time of the study. The participants of this study are part of a larger register study entitled PSY-PWS which takes place at Hannover Medical School and is described in more detail in former publications (28,29). The study followed the Declaration of Helsinki and was reviewed and approved by the local ethics committee of Hannover Medical School (No: 8129_BO_S_2020). All subjects or their legal representatives gave written informed consent to participation after all study procedures were explained to them.

The HQ-CT (Hyperphagia Questionnaire for Clinical Trials) in German language was used to assess the level of hyperphagia-related behavior in PWS. The HQ-CT is specifically designed and validated for PWS. It is a 9-item caregiver-report test instrument considered to be the current gold standard for hyperphagia assessment in PWS. The HQ-CT total score results from the sum of the 9 item-level responses ranging from 0 to 4 with a maximum score of 36 (30). Cut-off values above which clinically significant symptoms can be assumed have not yet been established.

### MOLECULAR ANALYSIS

DNA was extracted from blood samples using chemagicStar DNA-Blood1k kit (PerkinElmer Chemagen Technology, Baesweiler, Germany). The genomic DNA was subsequently bisulfite converted using the EpiTect® 96 Bisulfite Kit (Qiagen, Hilden, Germany) to detect the methylation status of the cytosine bases. Our target sequences within the LEP and LEPR promoter region were amplified from the previously purified bisulfite-converted DNA by polymerase chain reaction. Primers were designed to cover a LEP fragment of 823 bp (Hg 38, Chr. 7: 128.240.657 to 128.241.478), including exon 1 (Hg 38, Chr.7: 128.241.277 to 128.241.306), with 65 CpG-sites covered, and a LEPR fragment of 461 bp length (Hg 38, Chr. 1: 65.420.416 to 65.420.876), including exon 1 (Hg 38, Chr. 1: 65.420.651 to 65.420.741). The number of covered CpG-sites was 46.

Sequencing was performed using the Big-Dye® Terminator v3.1 Cycle Sequencing Kit (Applied Biosystems, Foster City, CA, USA). Individual PCR protocols and primer sequences can be found in **Supplementary Material S1**.

To determine the methylation status of the different CpG loci from the data obtained by DNA sequencing, Epigenetic Sequencing Methylation Analysis Software (ESME) was used (31). Methylation for each CpG site was calculated per each subject by building the ratio between normalized peak values of cytosine and thymine.

S-leptin levels were determined by enzyme-linked immunosorbent assay (ELISA) from Serum blood samples using Quantikine® Human Leptin Immunoassay (R&D Systems, Minneapolis, MN, USA).

### STATISTICS

Sequence Scanner v2.0 software (ABI Life Technologies, Grand Island, NY, USA) was used for sequence quality control. Only sequences showing a QV (quality value) > 20 were included for further analysis.

For statistical analysis, we used SPSS (IBM, Armonk, NY, USA). For graphical data processing, Graph Pad Prism (GraphPad, LaJolla, CA, USA) was used.

Data were further checked for missing values and variance. Single CpGs with more than 5% missing values and inter-individual variability below 5% were removed from the analysis. In addition, subjects with more than 5% missing values in methylation rate were excluded (see **Supplementary Material S2**).

Mann-Whitney test was used to compare differences in S-leptin levels and BMI between PWS and control subjects. We calculated mixed linear models to detect fixed effects of the variable group (PWS or control), CpG- position, sex and BMI on methylation rate. BMI and age were set as parameters of covariance. Bonferroni correction was applied to correct p-values for multiple testing. Differences in methylation rates at individual CpG sites were assessed within the mixed linear model with data splitted for CpG site. Fixed effects of factor group were calculated for the individual CpG sites. p values of < 0.05 were considered significant.

To assess the correlation between HQ-CT and the LEP/LEPR methylation in total and for individual CpG sites Pearson’s correlation coefficient was calculated. In addition, two groups were derived from the HQ-CT scores, with one group below and another group equal to or above the surveyed median HQ score of 8 in the PWS group for comparing LEP/LEPR methylation in pronounced hyperphagia with less pronounced hyperphagia. To this end, we examined effects of the factor “clinical severity of hyperphagia” on methylation rate according to the MLM detailed above and made group-wise comparisons.

## RESULTS

### PATIENT CHARACTERISTICS

The study included 24 subjects with Prader-Willi syndrome (male n=14, 58.3%; female n=10, 41.7%) and 13 healthy control subjects (male n=8, 61.5%; female n=5, 38.5%). In PWS subjects, the mean age was 27 years (minimum 12y, maximum 55y), whereas in controls, the mean age was 32 years (minimum 18y, maximum 47y). As matched for this, BMI values differed only slightly between groups showing no significant difference according to Mann Whitney test, U = 166, Z= 0.318, p = 0.766. Mean BMI was 27.2 kg/m^2^ in PWS (Minimum 19.1 kg/m^2^, Maximum 39.1 kg/m^2^) and 26.6 kg/m^2^ in healthy controls (Minimum 19.4 kg/m^2^, Maximum 40.0 kg/m^2^). Genetic subtype was determined in all subjects with PWS. DelPWS (n=11) and UPD (n=7) were shown most frequently (UPD/IC n=4 and IC n=2). Mean HQ-RT score in the PWS group was 10.39 (median 8, minimum 1, maximum 33).

**Table 1** presents the patient characteristics in detail.

**Table 1.**
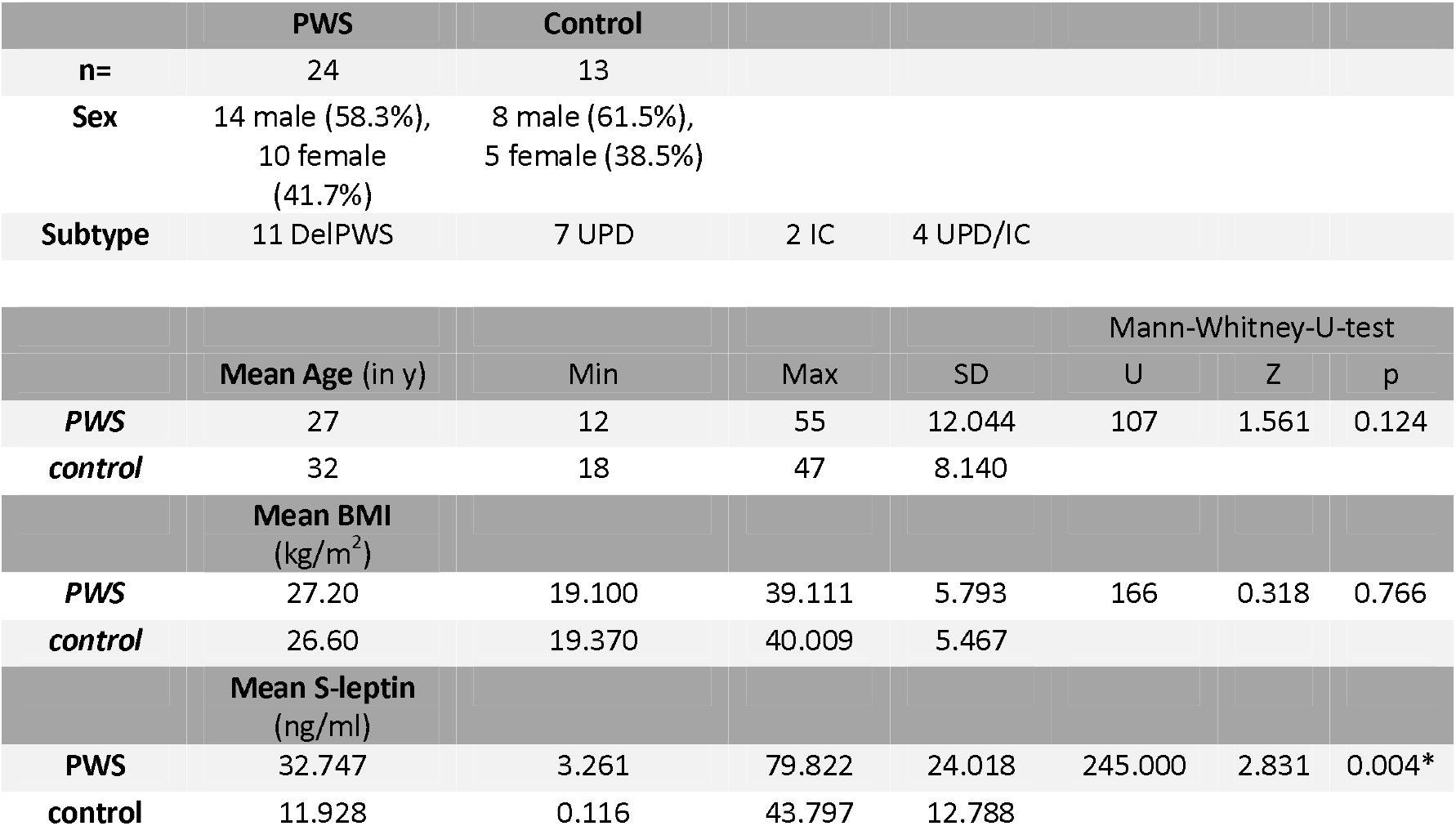
Demographics of PWS and control cohort

|  | PWS | Control |  |  |
| --- | --- | --- | --- | --- |
| **n=** | 24 | 13 |  |  |
| **Sex** | 14 male (58.3%),<br>10 female<br>(41.7%) | 8 male (61.5%),<br>5 female (38.5%) |  |  |
| **Subtype** | 11 DelPWS | 7 UPD | 2 IC | 4 UPD/IC |

**Table 1** Demographics of PWS and control cohort
|  | Mean Age (in y) | Min | Max | SD | Mann-Whitney-U-test |  |  |
| --- | --- | --- | --- | --- | --- | --- | --- |
|  |  |  |  |  | U | Z | p |
| **PWS** | 27 | 12 | 55 | 12.044 | 107 | 1.561 | 0.124 |
| **control** | 32 | 18 | 47 | 8.140 |  |  |  |
|  | Mean BMI<br>(kg/m<sup>2</sup>) |  |  |  |  |  |  |
| **PWS** | 27.20 | 19.100 | 39.111 | 5.793 | 166 | 0.318 | 0.766 |
| **control** | 26.60 | 19.370 | 40.009 | 5.467 |  |  |  |
|  | Mean S-leptin<br>(ng/ml) |  |  |  |  |  |  |
| **PWS** | 32.747 | 3.261 | 79.822 | 24.018 | 245.000 | 2.831 | 0.004* |
| **control** | 11.928 | 0.116 | 43.797 | 12.788 |  |  |  |

### S-LEPTIN

Mean S-Leptin in PWS subjects was 32.747 ng/ml (Minimum 3.261 ng/ml, Maximum 79.822 ng/ml) and 11.928 ng/ml in control group (Minimum 0.116 ng/ml, Maximum 43.797 ng/ml). Divided by sex, female subjects had higher serum leptin levels than males in both the PWS and control groups. Male PWS had a mean leptin level of 26,608 ng/ml (min 3,261, max 79,822), female of 41,316 ng/ml (min 14,755, max 68,826). Controls also showed a difference between males with an average of 5.324 ng/ml (Min 0.116, Max 16.213) to 22.493 ng/ml (Min 7.068, Max 43.797) in female controls.

According to Mann Whitney test for comparison of PWS and control groups in terms of S-leptin levels there was a significant difference between the groups, U = 245.00, Z= 2.831, p = 0.004 (see **Table 1** and **Figure 1**). Serum leptin levels correlate highly positively with BMI in both PWS subjects (linear R2 = 0.502) and healthy control subjects (linear R2 = 0.623). S-Leptin values do not correlate with the HQ results in the PWS subjects (linear R2 = 0.016).

**Figure 1.**
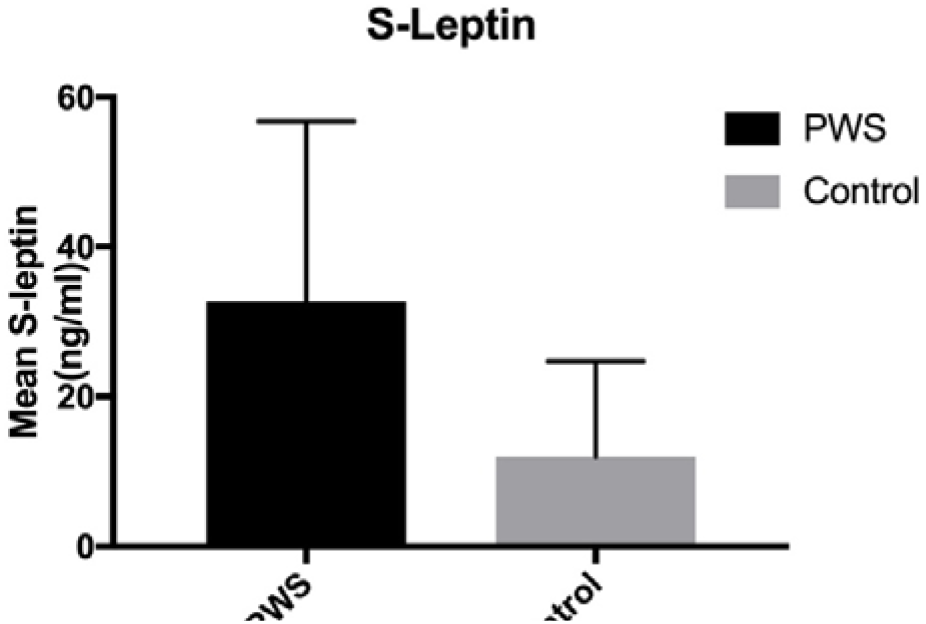
Chart showing significant higher mean S-Leptin (with SD) in PWS compared to BMI-matched controls.

### LEP METHYLATION

For LEP significant fixed effects on methylation rate of individual CpG position (F_(48,1660)_ = 85,312; p < 0.001) and group (F _(1,1660)_ = 22,706, p < 0.001) were detected. MLM revealed no significant CpG x group interaction for LEP gene (F _(48,1660)_ = 1,247; p = 0,121). We found independent effects of age (F _(1,1660)_ = 34,185; p < 0,001), but not of sex (F_(1, 1660)_ = 2,657, p = 0,103) nor BMI (F_(1, 1660)_ = 1,52, p = 0,218). Significance of group differences was not affected when age and gender were applied as additional factors.

Overall LEP mean methylation rate was shown to be lower in PWS (29.9%) compared to healthy controls (32,5%) (see **Figure 2**). Analysis of fixed effects of factor group on single CpG sites methylation revealed significant effects for eleven individual CpG positions (m169, m199, m203, m329, m331, m338, m340, m349, m371, m373 and m378). All CpG sites named above showed a significantly lower mean methylation rate in the PWS group compared to controls. The CpG positions m329 and m331, m338 and m 340, and m349 to m373 sequentially follow each other in the base sequence. **Figure 3** shows a graphical representation of these results. No significant differences in mean methylation rate were found between the sexes with males showing an insignificantly higher methylation rate (31,6%) compared to females (30,8%). For detailed statistical results see **Supplementary Material S3**.

**Figure 2.**
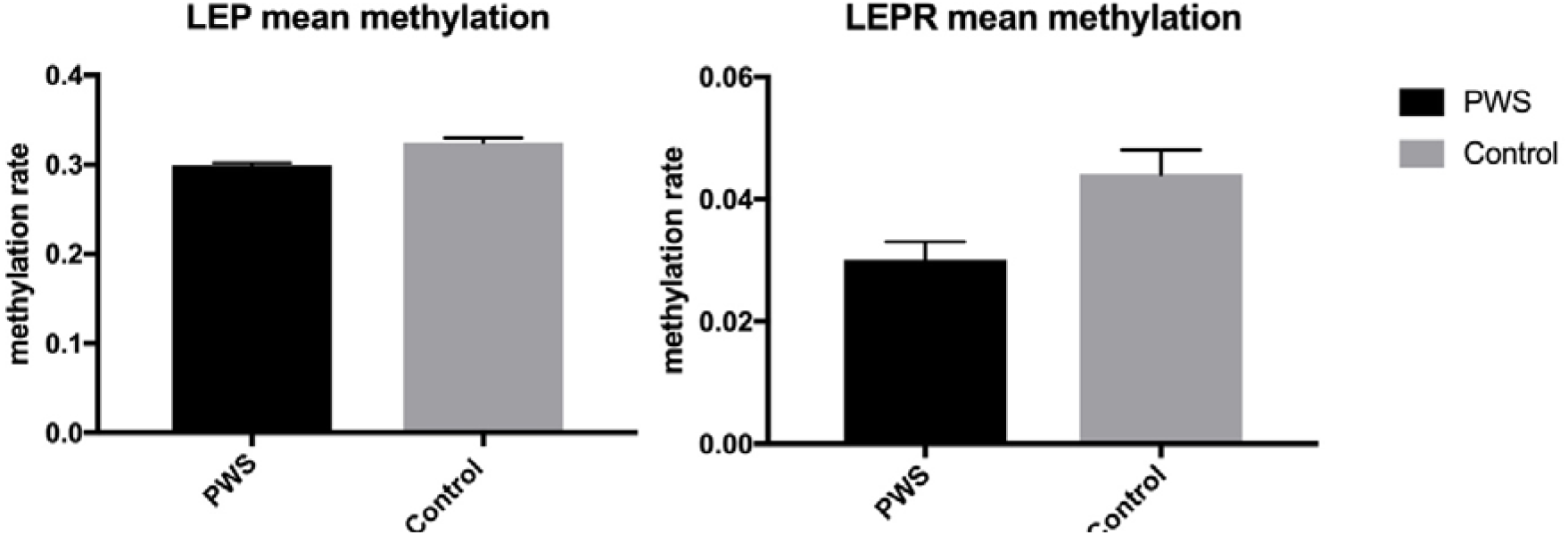
Chart showing significant lower mean LEP and LEPR promoter methylation (with SD) in PWS compared to BMI-matched controls.

**Figure 3.**
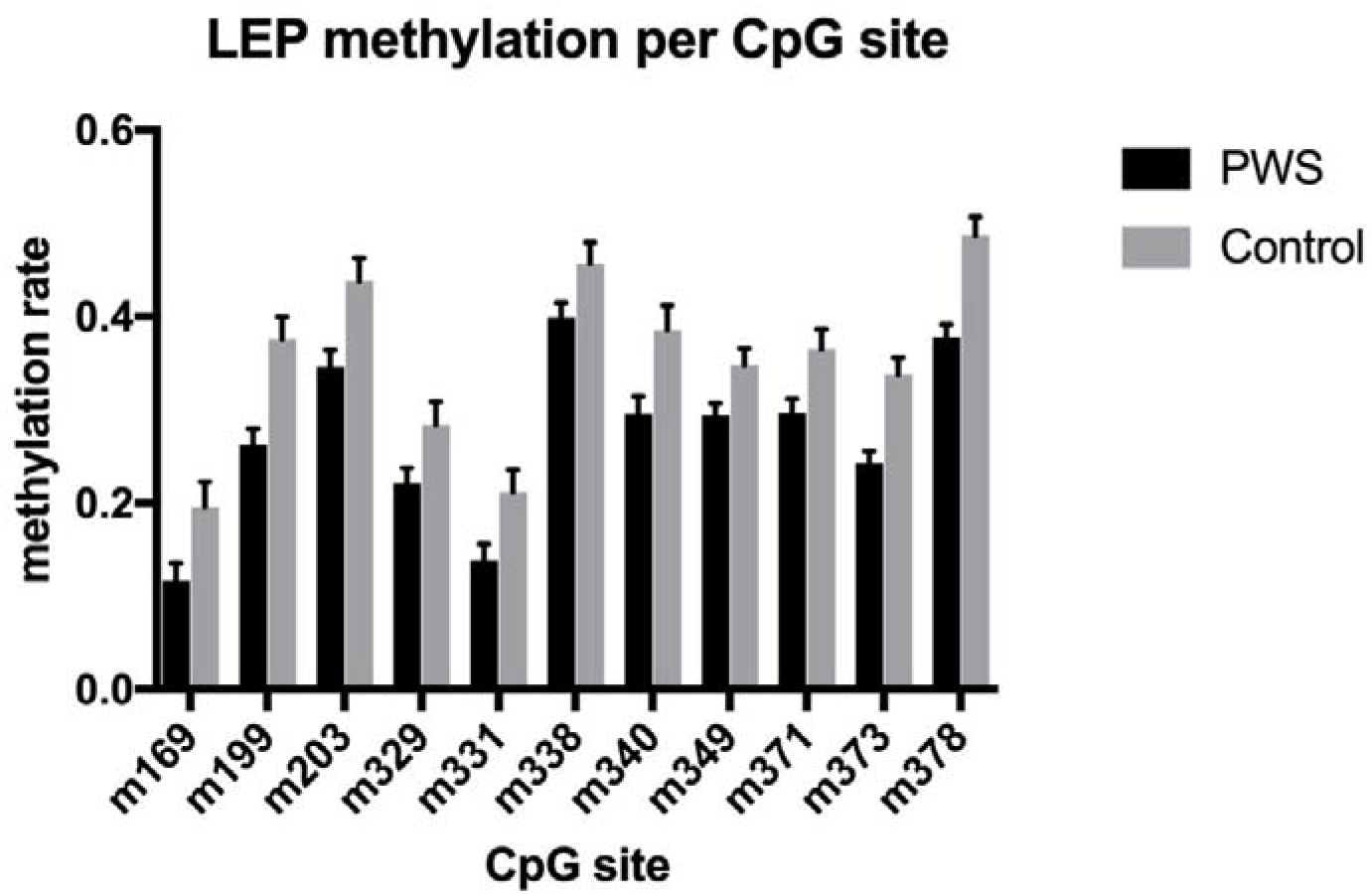
Chart revealing significant lower mean methylation of LEP-promoter (with SD) in eleven of partially contiguous CpG regions in PWS compared to controls (only shown CpG sites with a significant difference)

Correlation analysis revealed no significant correlation between overall LEP promoter methylation rate and S-Leptin levels. Also, there was no correlation between overall LEP methylation and HQ scores.

According to Pearson’s r, there was a negative correlation between mean methylation at CpG sites m349 (r = - 0.473, p = 0.023) and m371 (r = - 0,462, p = 0,026) and HQ scores in subjects with PWS. No significant correlation was found for any other LEP CpG site shown to differ significantly between the groups in LEP methylation rate. The MLM showed no significant effect of the factor “clinical severity of hyperphagia” on the methylation rate (F _(1,1076)_ = 1.571; p = 0.210).

### LEPR METHYLATION

MLM revealed significant fixed effects of factors group (F_(1,813)_ = 11,021; p = 0.001) as well as CpG-position (F_(23,813)_ = 8,984; p < 0.001) on the LEPR methylation rate. Overall LEPR mean methylation rate was lower in PWS with 3.0% compared to 4.4% in controls. In contrast to LEP analysis, we showed a significant group x CpG site interaction for LEPR gene (F_(23,813)_ = 2,414; p < 0.001). No significant fixed effects were seen for factor sex on methylation rate (p = 0.279), nor for age (p = 0.977) or BMI (p = 0.382). Significance of group differences was not affected when age and gender were applied as additional factors.

Within the mixed linear model analysis split for single CpG sites, significant effects of factor group at eight individual CpG positions were seen (m211, m196, m86, m4, p31, p76, p88 and p129). No contiguous CpG positions could be identified. **Figure 4** shows a graphical representation of these results. For detailed statistical results see **Supplementary Material S4**.

**Figure 4.**
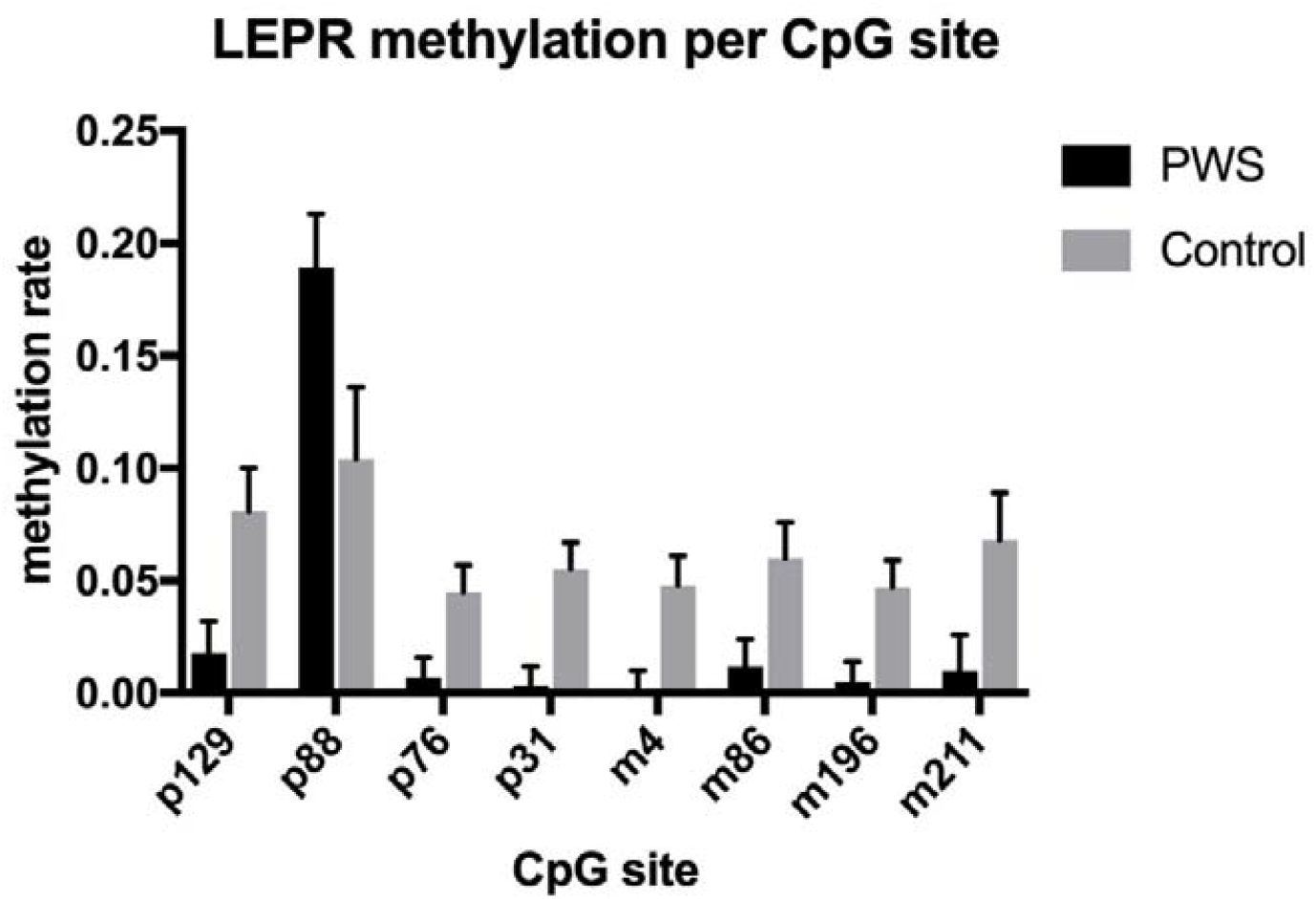
Chart revealing significant difference in mean methylation of LEPR-promoter (with SD) in eight CpG regions in comparison of PWS and controls (only shown CpG sites with a significant difference)

Correlation analyses showed a highly significant inverse correlation between overall LEPR methylation rate and S-Leptin levels with Pearsons r = -0,120, p < 0,001. No correlation was found between overall LEPR methylation rate and HQ scores. There were no correlations of individual CpG sites methylation rate with HQ scores. The MLM showed no significant effect of the factor “clinical severity of hyperphagia” on the methylation rate (F_(1,503)_ = 0.018; p = 0.893).

## DISCUSSION

The present study investigates serum leptin levels and methylation of the LEP and LEPR promoter in adult PWS compared to healthy BMI-matched controls. Our study reveals that PWS subjects have significantly higher S-leptin levels than controls while showing significantly downregulated LEP and LEPR promoter methylation. LEPR, but not LEP methylation correlated with S-leptin levels, while overall there was no correlation with the behavioral dimension in terms of the HQ-scores.

To our knowledge, the study is the first to demonstrate a significant difference in serum leptin levels between obese adults with PWS and weight-matched controls, a finding previously shown only for infants <6 years of age that appeared to resolve with increasing age. In line with previous studies a correlation of leptin level with BMI was still observed in both PWS and control group in the present study, but as they were matched for BMI both the PWS and control group were only slightly overweight at 27.2 respective 26.6 kg/m^2^. The mean BMI of our study seems to be considerably lower compared to other studies on the topic, for example, 37.2 kg/m^2^ in Butler et al. or 41.2 kg/m^2^ in Pagano et al. (19,20). Altogether, a BMI-independent effect of the factor PWS on S-leptin levels can be assumed. The comparatively high number of male subjects in our study might have a confounding effect, especially since sex-dependent differences in leptin regulation are known. An indication could be the study by Butler et al., which showed comparatively higher leptin blood levels only in male PWS subjects, who, unlike in our study, were non-obese (20). But this idea again conflicts with previous findings that women generally have higher leptin levels than men. In contrast, Kennedy et al. also showed no differences in leptin levels between PWS and overweight controls, although females significantly predominated in the PWS cohort (18). Our results displayed no significant differences between the sexes in terms of blood leptin levels.

Furthermore, the present study is the first to investigate the epigenetic regulation of the satiety hormone leptin in people with PWS. In terms of obesity in general the majority of previous human studies have demonstrated an association between decreased LEP methylation levels and elevated leptin serum levels in obese subjects, hence an inverse correlation between LEP promoter methylation and leptin expression was assumed. Our results appear partly conclusive on this point from a neurobiological perspective, although it should be mentioned that no significant correlation was shown between LEP promoter methylation and S-leptin levels. Also, Lindgren et al. found no difference in leptin mRNA levels between PWS and obese controls, although increased leptin protein biosynthesis might be expected with decreased LEP promoter methylation as found in the present cohort (21). Also, data from our laboratory regarding LEP promoter methylation after bariatric surgery recently indicated decreased LEP promoter methylation with concomitant decreased serum leptin after surgically induced weight loss, contrary to the majority of observations made before (26). Overall, however, intervention-induced weight loss with the accompanying hormonal disruptions appears to bear little resemblance to the given conditions in PWS. In contrast, analogous to the study by Wilhelm et al, we also see an inverse correlation between LEPR methylation and S-leptin levels in the present cohort. Taken together, the findings of hypomethylated LEP/LEPR promoter and elevated S-leptin made in our PWS cohort mirror those previously made in obesity in general.

With central leptin resistance known to occur in obesity and suspected in PWS, a previous study suggested that above a count of 25-30ng/ml S-leptin central leptin resistance can be assumed (32), a limit which is exceeded by our PWS cohort. Given the abovementioned similarities of our findings in PWS to obesity in general, could PWS even serve as a model disease for leptin resistance?

The following hypothesis appears to emerge from the evidence to date. Among the chromosomal region affected in PWS is the melanoma antigen gene family member L2 (MAGEL2) gene. It is known that leptin-mediated activation of anorexigenic POMC neurons in the arcuate nucleus of the hypothalamus is mediated via leptin receptors LepRb. Leptin-induced POMC-activation was absent in a Magel2 knockout mouse model, suggesting that MAGEL2 is required for the leptin-mediated responses that is critical for POMC neuron-mediated suppression of food intake (33). To that end, it can be hypothesized that loss of Magel2 disrupts the normal balance of LepR cell surface expression, because MAGEL2 and Necdin, another gene encoded in the PWS region, regulate the linkage of leptin receptors to an ubiquitination complex relevant for surface expression, internalization, and degradation processes. In contrast, while MAGEL2 is normally associated with cell surface abundance of LepR and its reduced degradation, the hypothalamus of Magel2-null mice showed less LepR and altered levels of proteins of the ubiquitin pathway that regulate LepR processing (34). Hypothalamic LepRbs are generally suspected to be involved in the development of leptin resistance. Downregulation of LepRb mRNA and proteins and impaired transport of LepRb to the plasma membrane in neuronal subpopulations of hypothalamic nuclei that control energy homeostasis have emerged as possible molecular mechanisms of leptin resistance before (35). Limited by the fact that we only investigated in peripheral blood DNA-methylation, our observation of significantly elevated serum leptin levels with decreased LEP and LEPR promoter methylation could represent a counter regulatory mechanism in response to leptin resistance in PWS genetically determined by MAGEL2 and Necdin derived downregulated LepRb expression in the hypothalamus satiety center. We hypothesize that central leptin resistance is counteracted by upregulated leptin hormone and leptin receptor expression in PWS mediated at the epigenetic level via decreased promoter methylation of LEP and LEPR, as we have seen analogously in previous studies on human obesity showing an inverse correlation between LEPR methylation and LepRb expression, for example (27). We see corroboration of our theory by our observation that LEPR, but not LEP promoter methylation, correlated significantly with S-leptin levels in the subjects, suggesting a functional regulatory link for LEPR. However, it remains to be mentioned that Wijesuriya et al. also noted that Magel2 mutation rodent models showed an overall less pronounced hyperphagia phenotype than other models, for example compared to Snord116 gene deficiency.

Likewise, our study failed to provide a conclusive link between laboratory findings and the behavioral dimension. Overall, no correlation of serum leptin levels with HQ scores was demonstrated in the PWS subjects. Also, no correlation was found between overall methylation rate of LEP and LEPR with HQ-scores in PWS, however solely for LEP CpG sites m349 and m371. From a neurobiological perspective, our finding that the methylation rates of the abovementioned CpG sites correlate negatively with the outcome of the hyperphagia questionnaire appears inconclusive. As stated above, it should be assumed that increased leptin expression via decreased LEP methylation should be accompanied by an increased feeling of satiety and, accordingly, by decreased food-seeking behavior, which would correspond to a lower score on the HQ. In this respect, the correlations found seem most likely to be incidental. To this end, our findings appear to be in line with previous human studies on this topic like Goldstone et al. being unable to establish a connection between hormonal abnormalities in PWS and the behavioral dimension in the sense of an altered eating behavior (14). In synopsis of our and previous findings, no connection between leptin regulation and behavioral abnormalities in PWS can be assumed.

### LIMITATIONS

Overall, the cohort size of our study is small, but in line with that of previous studies on the topic.

Especially limiting appears the use of peripheral blood material, although the regulation of satiety is essentially due to central processes. Therefore, only indirect conclusions can be drawn regarding the central processes behind leptin resistance in PWS. Unfortunately, we did not have central material such as CSF available for our study, as such sampling is hardly justifiable from an ethical point of view.

Furthermore, the HQ currently is described to be the gold standard for assessing food-seeking behavior in PWS. However, the overall data on the validity of the questionnaire appears limited. In general, it offers only an indirect assessment, since it is completed by relatives or caregivers, which could be subject to perceptual bias compared to the PWS individuals themselves. Also, the HQ was not performed in the control subjects, so comparability is not given.

Another crucial limitation of the study seems to be the use of BMI as determinant of obesity in the subjects. BMI values of the control group could likely be confounded by a comparatively increased muscle mass. Body fat percentage could be the much more accurate determinant for comparison at this point, especially since leptin is primarily produced in adipocytes. However, the comparability with previous studies still appears to be given insofar as these also mostly used body weight or BMI as assessment standard.

Furthermore, our study lacks a healthy, normal-weight control group for comparisons regarding leptin resistance in obese and non-obese subjects.

### CONCLUSION

The observed differences in leptin expression again pointed towards a dysregulation of the hormonal network regulating hunger and satiety in people with PWS without being able to provide a detectable connection of leptin dysregulation and food-seeking behavior typical in PWS. But overall, our research findings strengthen the hypothesis that PWS could be considered as a model disease for leptin resistance and thus be of great interest for obesity research in general.

## Data Availability

All data produced in the present study are available upon reasonable request to the authors.

## AUTHOR CONTRIBUTION

Jelte Wieting contributed in acquisition of samples, drafted the manuscript and contributed to analysis and interpretation of data. Kirsten Jahn carried out the experiments and contributed to analysis and interpretation of data. Vanessa Buchholz and Ralf Lichtinghagen carried out the experiments. Christian K. Eberlein contributed to acquisition of samples and contributed to interpretation of data and revised the manuscript critically for important intellectual content. Stefan Bleich contributed to interpretation of data and revised the manuscript critically for important intellectual content. Maximilian Deest contributed to acquisition of samples, analysis, and interpretation of the data and was substantially involved in drafting the manuscript. Helge Frieling contributed to conception and design of the study, interpretation of data, and revised the manuscript critically for important intellectual content.

