## Supplementary Material for "Alteration of serum leptin and LEP/LEPR promoter methylation in Prader-Willi syndrome"

**S1**

MOLECULAR ANALYSIS

LEP PCR

Samples were incubated at 95°C for 15 minutes and 97 °C for 1 minute. Subsequently 15 cycles composed of the following three steps: 95°C for 30 seconds, 72 °C for 45 seconds and 68 °C for 1 minute were applied, followed by 25 cycles of 95 °C for 30 seconds, 57 °C for 45 seconds and 65 °C for 1 min 30 sec. Finally, 65 °C were applied for 5 minutes and samples were hold at 12 °C until further processing. The primer sequences used for LEP PCR are TGGGGTGTTAGTTAGAGAT (738_LEP_F1_neu) and ATAACCTTCTATCTAACTAAAAC (739_LEP_R1_neu) (IDT®).

LEPR PCR

samples were incubated at 95°C for 15 min followed by 5 cycles composed of the three steps 95°C for 30 sec, 65°C for 1 min 30 sec and 72°C for 2 min 30 sec. Subsequently 28 cycles of the three steps 95°C for 30 sec, 65°C for 45 sec and 72°C for 1 min were applied. Finally, 72°C were applied for 4 min and samples were hold at 12°C until further processing. The primer sequences used for LEPR PCR are GGATTAGTAGGGGAGGTTTT (374-LEPR_T2_F1) and AAAATAACAACCCCACCACA (377-LEPR_T2_R2).

Both were performed on a C1000 Thermal Cycler (BIO-RAD, Hercules, CA, USA) using the HotStarTaq® Master Mix Kit (Qiagen, Hilden, Germany). PCR product was purified automatically on a Biomek® NxP (Beckman Coulter, Brea, CA, USA) using paramagnetic beads (Clean-NGS, GC Biotech®).

SEQUENCING PCR

Sequencing was performed using the Big-Dye® Terminator v3.1 Cycle Sequencing Kit (Applied Biosystems, Foster City, CA, USA). The forward primer (738_LEP_Forward) was used for LEP sequencing PCR.

The PCR protocol (SeqATRev) implied 96 °C for 1 minute, followed by 25 cycles composed of 96 °C for 5 seconds, 60 °C for 90 seconds and 50 °C for 90 seconds. Forward primer (374-LEPR_T2_F1) was used for LEPR sequencing PCR. The PCR protocol (SeqSTD) implied 96°C for 1 min, followed by 28 cycles composed of 96°C for 10 sec, 50°C for 5 sec and 60°C for 4 min. Samples were Both samples were subsequently hold at 12°C until further processing.

Purification of sequencing PCR was automized via Biomek NxP using CleanDTR beads (CG Biotech®). Sequencing itself was performed on a 3500XL genetic analyzer (ABI Life Technologies, Grand Island, NY, USA).

**S2**

LEP

A total of 13 out of the 65CpGs were excluded from the analysis during quality control, n=8 (m187, m32, m0, p20, p9, p39, p42, p44, p14) due to missing values and n=5 (m117, m61, m70, m73, m84) due to low variance. Control subject ID09 was excluded due to missing LEP methylation values.

LEPR

A total of 22 CpGs were excluded from further analysis, n=12 (m106, p111, p116, p139, p141, p164, p168, 182, p184, p56, p61, p67) due to missing values and n=10 (m112, m162, m171, m203, m44, m66, m88, p152, p74, p81) due to low variance. No subject was excluded due to missing values.

**S3**

| **Test for fixed effects, type IIIa** | | |  |
| --- | --- | --- | --- |
| **Factor** | ***df*** | ***F*** | ***Sig.*** |
| ***group*** | 1 , 1660 | 22,706 | <0,001* |
| ***CpG*** | 48 , 1660 | 85,312 | <0,001* |
| ***sex*** | 1 , 1660 | 2,657 | 0,103 |
| ***age*** | 1 , 1660 | 34,185 | <0,001* |
| ***BMI*** | 1 , 1660 | 1,52 | 0,218 |
| **group * CpG** | 48 , 1660 | 1,247 | 0,121 |

| **Estimates** |  |  |  |  |  |
| --- | --- | --- | --- | --- | --- |
| ***Group*** | ***Mean*** | **SD** | **df** | **Lower 95%CI** | **Upper 95%CI** |
| *control* | 0,325 | 0,005 | 1708 | 0,317 | 0,334 |
| *PWS* | 0,299 | 0,003 | 1708 | 0,293 | 0,305 |
| ***Sex*** |  |  |  |  |  |
| *m* | 0,316 | 0,003 | 1708 | 0,31 | 0,323 |
| *f* | 0,308 | 0,004 | 1708 | 0,3 | 0,316 |
| Dependent variable: methylation rate. | | | |  |  |
| The covariates in the model are calculated using the following values: Age = 29.01, BMI = 27.174237717893. | | | | | |

| **CpG** | **Fixed Effect** | | | **Estimates** | |  |  |  |
| --- | --- | --- | --- | --- | --- | --- | --- | --- |
|  | **df** | **F-value** | **Sig.** |  | **Mean** | **SD** | ***Lower***  ***CI95*** | ***Upper***  ***CI 95*** |
| **m169** | 1,34 | 5,686 | 0,023 | *Control* | 0,196 | 0,027 | 0,141 | 0,251 |
|  |  |  |  | *PWS* | 0,117 | 0,019 | 0,078 | 0,156 |
| **m199** | 1,34 | 14,536 | 0,001 | *Control* | 0,376 | 0,024 | 0,327 | 0,425 |
|  |  |  |  | *PWS* | 0,263 | 0,017 | 0,228 | 0,298 |
| **m203** | 1,34 | 9,164 | 0,005 | *Control* | 0,438 | 0,025 | 0,388 | 0,489 |
|  |  |  |  | *PWS* | 0,346 | 0,018 | 0,31 | 0,382 |
| **m329** | 1,34 | 4,381 | 0,044 | *Control* | 0,284 | 0,025 | 0,234 | 0,334 |
|  |  |  |  | *PWS* | 0,221 | 0,017 | 0,186 | 0,257 |
| **m331** | 1,34 | 5,988 | 0,02 | *Control* | 0,212 | 0,024 | 0,163 | 0,261 |
|  |  |  |  | *PWS* | 0,139 | 0,017 | 0,104 | 0,174 |
| **m338** | 1,34 | 4,121 | 0,05 | *Control* | 0,457 | 0,023 | 0,41 | 0,504 |
|  |  |  |  | *PWS* | 0,399 | 0,016 | 0,366 | 0,432 |
| **m340** | 1,34 | 7,717 | 0,009 | *Control* | 0,386 | 0,026 | 0,332 | 0,439 |
|  |  |  |  | *PWS* | 0,296 | 0,019 | 0,258 | 0,334 |
| **m349** | 1,34 | 5,963 | 0,02 | *Control* | 0,348 | 0,018 | 0,311 | 0,384 |
|  |  |  |  | *PWS* | 0,294 | 0,013 | 0,268 | 0,32 |
| **m371** | 1,34 | 6,39 | 0,016 | *Control* | 0,365 | 0,022 | 0,321 | 0,409 |
|  |  |  |  | *PWS* | 0,297 | 0,015 | 0,266 | 0,329 |
| **m373** | 1,34 | 18,82 | <0.001 | *Control* | 0,338 | 0,018 | 0,301 | 0,374 |
|  |  |  |  | *PWS* | 0,243 | 0,013 | 0,218 | 0,269 |
| **m378** | 1,34 | 19.923 | <0.001 | *Control* | 0,487 | 0,02 | 0,446 | 0,527 |
|  |  |  |  | *PWS* | 0,378 | 0,014 | 0,349 | 0,407 |

**S1** Mixed linear model LEP gene: Listed are the fixed effects of the above factors on methylation rate and the group comparison of PWS and controls and sexes. Further shown are the effects of the factor group on the methylation of individual CpG positions with pairwise comparison, only those with significant group effect are mentioned.

**S4**

| **Test for fixed effects, type IIIa** | | | |
| --- | --- | --- | --- |
|  | ***df*** | ***F*** | ***sig.*** |
| ***group*** | 1 , 813 | 11,021 | 0,001* |
| ***CpG*** | 23 , 813 | 8,984 | <0,001* |
| ***sex*** | 1 , 813 | 1,175 | 0,279 |
| ***age*** | 1 , 813 | 0,001 | 0,977 |
| ***BMI*** | 1 , 813 | 0,764 | 0,382 |
| ***group * CpG*** | 23 , 813 | 2,414 | <0,001* |

| **Estimates** |  |  |  |  |  |
| --- | --- | --- | --- | --- | --- |
| ***Group*** | ***mean*** | **SD** | **df** | **Lower 95%CI** | **Upper95%CI** |
| *control* | ,044 | 0,004 | 836 | 0,037 | 0,05 |
| *PWS* | ,030 | 0,003 | 836 | 0,025 | 0,04 |
| ***Sex*** |  |  |  |  |  |
| *m* | ,039 | 0,003 | 836 | 0,034 | 0,05 |
| *f* | ,035 | 0,003 | 836 | 0,028 | 0,04 |
| Dependent variable: methylation rate. | | | | | |
| The covariates in the model are calculated using the following values: BMI = 27.021647174828, age = 29.06. | | | | | |

| **CpG** | **Fixed Effect** |  |  | **Estimates** |  |  |  |  |
| --- | --- | --- | --- | --- | --- | --- | --- | --- |
|  | **df** | **F-value** | **Sig.** |  | **Mean** | **SD** | ***Lower CI95*** | ***Upper CI 95*** |
| **p129** | 1,34 | 7,033 | 0,012 | *Control* | 0,081 | 0,019 | 0,042 | 0,119 |
|  |  |  |  | *PWS* | 0,018 | 0,014 | -0,011 | 0,047 |
| **p88** | 1,34 | 4,517 | 0,041 | *Control* | 0,104 | 0,032 | 0,039 | 0,169 |
|  |  |  |  | *PWS* | 0,189 | 0,024 | 0,14 | 0,238 |
| **p76** | 1,34 | 6,15 | 0,018 | *Control* | 0,045 | 0,012 | 0,02 | 0,071 |
|  |  |  |  | *PWS* | 0,007 | 0,009 | -0,012 | 0,026 |
| **p31** | 1,34 | 11,423 | 0,002 | *Control* | 0,055 | 0,012 | 0,03 | 0,079 |
|  |  |  |  | *PWS* | 0,003 | 0,009 | -0,015 | 0,022 |
| **m4** | 1,34 | 8,515 | 0,006 | *Control* | 0,048 | 0,013 | 0,021 | 0,074 |
|  |  |  |  | *PWS* | 0 | 0,01 | -0,02 | 0,02 |
| **m86** | 1,34 | 5,992 | 0,02 | *Control* | 0,06 | 0,016 | 0,028 | 0,092 |
|  |  |  |  | *PWS* | 0,012 | 0,012 | -0,012 | 0,036 |
| **m196** | 1,34 | 7,592 | 0,009 | *Control* | 0,047 | 0,012 | 0,022 | 0,072 |
|  |  |  |  | *PWS* | 0,005 | 0,009 | -0,014 | 0,023 |
| **m211** | 1,34 | 5,089 | 0,031 | *Control* | 0,068 | 0,021 | 0,026 | 0,111 |
|  |  |  |  | *PWS* | 0,01 | 0,016 | -0,022 | 0,041 |

**S2** Mixed linear model LEPR gene: Listed are the fixed effects of the above factors on methylation rate and the group comparison of PWS and controls and sexes. Further shown are the effects of the factor group on the methylation of individual CpG positions with pairwise comparison, only those with significant group effect are mentioned.
